# Optimal Control applied to a SEIR model of 2019-nCoV with social distancing

**DOI:** 10.1101/2020.04.10.20061069

**Authors:** Abhishek Mallela

## Abstract

Does the implementation of social distancing measures have merit in controlling the spread of the novel coronavirus? In this study, we develop a mathematical model to explore the effects of social distancing on new disease infections. Mathematical analyses of our model indicate that successful eradication of the disease is strongly dependent on the chosen preventive measure. Numerical computations of the model solution demonstrate that the ability to flatten the curve becomes easier as social distancing is strictly enforced. Based on our model, we also formulate an optimal control problem and solve it using Pontryagin’s Maximum Principle and an efficient numerical iterative method. Our numerical results of an optimal 2019-nCoV treatment protocol that yields a minimum disease burden from this disease indicates that social distancing is vitally important.

## 1 Introduction

The novel coronavirus 2019-nCoV has been spreading around the globe at a frightening pace. Academic institutions around the world have canceled in-person instruction, traveling between places is strongly discouraged, and organizations have endorsed teleworking policies. These efforts are collectively referred to as *social distancing*. The main idea behind these efforts is to reduce human contact in order to reduce the risk of infectivity. How can we assess the value of such strategies and model the spread of the disease?

Mathematical models are a useful tool to investigate disease dynamics. Existing models on coronavirus have provided greater insights into the transmission dynamics, the prevention of new infections, and methods of parameter estimation [3, 5, 7]. However, none of these models have explored a viable social distancing strategy using optimal control theory. In this study we develop an epidemiological model that assesses the value of social distancing strategies. Using our model, we also formulate an optimal control problem to identify the optimal treatment protocol that provides a minimum disease burden from the coronavirus.

The paper is organized as follows: in Section 2, we formulate the SEIR model. In Section 3, we conduct mathematical analyses, including the formulation of the basic reproduction number and the stability analysis of the disease-free equilibrium. In Section 4, numerical computations of the model solution are performed to explore the effects of social distancing. In Section 5, we formulate an optimal control problem, provide a method of numerical computation, and discuss optimal control results. We summarize our findings with conclusion in Section 6.

## 2 Model Description

To address the key question of this study, we use the *SEIR* compartmental model framework, where *S, E, I* and *R* denote the susceptible, exposed, infected, and recovered populations respectively. A schematic diagram of the model is given in Fig. 1. Upon introduction of the disease, individuals transition among these compartments. For the sake of simplicity, we assume that a recovered individual is no longer able to be infected. The incubation frequency of the virus is denoted by *α* and the inverse of the average length of infection is given by *γ*. The parameter *β* is defined as the average rate of person-to-person contact times the infectivity (the probability per that an infected individual transmits the disease to a susceptible individual). We account for the effect of social distancing with the parameter *σ*. Social distancing refers to the absence of large gatherings, avoidance of physical contact, and other efforts necessary to mitigate the spread of 2019-nCoV.

**Figure 1:**
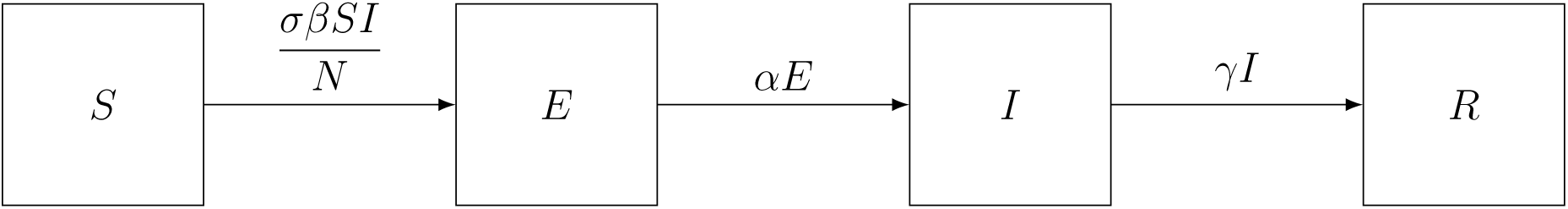
Schematic diagram of the model. The box compartments represent groups of individuals and the arrows represent disease transmission, disease incubation, and disease progression. *S* = susceptible, *E* = exposed, *I* = infected, *R* = recovered

Thus, we can write the model mathematically as:

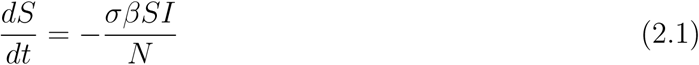

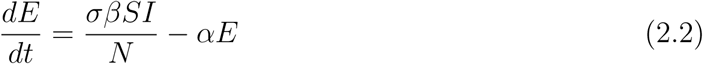

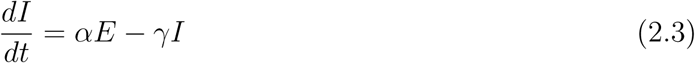

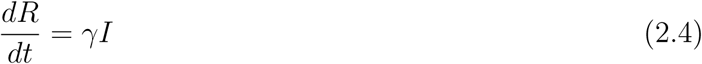

where the total population is *N* (*t*) = *S*(*t*) + *E*(*t*) + *I*(*t*) + *R*(*t*).

Using our model, we can compute the total number of new infections *T*_*N*_ as follows:

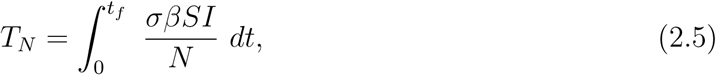

where *t*_*f*_ denotes the final time of the study period.

The number of new infections is a major concern of 2019-nCoV epidemiology and treatment. Therefore, we take a simple approach and define the total disease burden for the fixed time interval, [0, *t*_*f*_], as *A*_1_*T*_*N*_, where *A*_1_ is a weighting coefficient.

## 3 Model Analysis

The variables in the model system (2.1) - (2.4) remain non-negative for all *t ≥* 0 with non-negative initial conditions, because 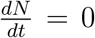 implies that *N* is a non-negative constant (in order for the system to be biologically meaningful).

### 3.1 Basic Reproduction Number

The basic reproduction number, ℛ_0_, is defined as the effective number of secondary infections caused by a typical infected individual during his/her entire period of infectiousness, when he/she is introduced into a population consisting of susceptible individuals only. Here, we formulate ℛ_0_ using the next-generation matrix method [1, 9]. The model has a unique disease-free equilibrium (DFE): *E*_0_ = (*N*, 0, 0, 0). For the next-generation method, we consider equations corresponding to the infectious compartments only (i.e. *E* and *I*). The non-negative matrix, ℱ, corresponding to the new infections in the population, evaluated at the disease-free equilibrium *E*_0_, is given by:

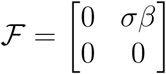

The nonsingular matrix, *𝒱*, corresponding to the transfer of individuals in and out of compartments, is:

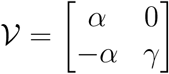

Then ℛ_0_, which corresponds to the dominant eigenvalue of the matrix ℱ *𝒱*^*−*1^, is given by

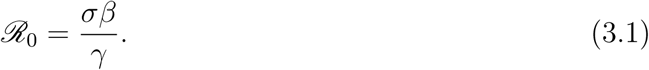

### 3.2 Stability Analysis of Disease-Free Equilibrium

From results in [9], ℛ_0_ provides the local stability criteria for the DFE:

#### Proposition 3.2.1.

*The DFE of* (2.1) *-* (2.4) *is locally asymptotically stable if* ℛ_0_ < 1 *and is unstable if* ℛ_0_ > 1.

We are also able to prove that ℛ_0_ < 1 implies global stability of the DFE as shown in the following theorem:

#### Theorem 3.2.2.

*The DFE of* (2.1) *-* (2.4) *is globally asymptotically stable if* ℛ_0_ < 1.

*Proof:* Let *I*^*C*^ denote the total number of infected individuals, i.e., *I*^*C*^ = *E* + *I*. Then,

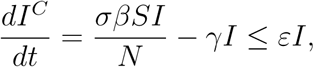

where *ε* = (*σβ − γ*). If ℛ_0_ < 1, then *ε* < 0. This implies that the solution curve *I*^*C*^(*t*) is bounded above by a solution that decays exponentially to zero if ℛ_0_ < 1. Therefore, if ℛ_0_ < 1 and (*E, I*) (0, 0), then *I*^*C*^(*t*) decreases and eventually approaches zero as *t → ∞*. Since *E, I ≥* 0, both *E* and *I* approach zero in the limit as *t → ∞*. This implies that

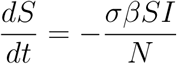

approaches 0 as *t → ∞*. Thus we have:

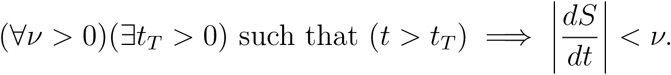

Thus, we can conclude that lim_*t→∞*_*S*(*t*) = *N*. Hence, the disease-free equilibrium is globally asymptotically stable if ℛ_0_ < 1.

## 4 Model Predictions

### 4.1 Parameter Estimation

We estimated the model parameters based on the literature survey [3, 5, 7]. The parameters used for our simulations are given in Table 1. For our model predictions, we focus on the total new infections, *T*_*N*_. For the sake of demonstration, we compute the totals over a period of 200 days, i.e., *t*_*f*_ = 200 days. Since we do not know the value for the weightage coefficient, for simplicity, we take *A*_1_ = 1 for our model simulations.

**Table 1:** Model parameters

| Description | Entity | Estimate | Source |
| --- | --- | --- | --- |
| Total population | N | 10,000 | Assumed |
| **INITIAL CONDITIONS** |  |  |  |
| Susceptible | $S(0)$ | 9,999 | Assumed |
| Exposed | $E(0)$ | 0 | Assumed |
| Infected | $I(0)$ | 1 | Assumed |
| Recovered | $R(0)$ | 0 | Assumed |
| **PARAMETERS** |  |  |  |
| Incubation rate | $\alpha$ | $1.818 \times 10^{-1} \text{ day}^{-1}$ | [3] |
| Mean contact rate | $\beta$ | $1.68 \text{ day}^{-1}$ | [5] |
| Mean infection rate | $\gamma$ | $5 \times 10^{-1} \text{ day}^{-1}$ | [7] |
| Coefficient of social distancing | $\sigma$ | 0 to 1 | Varied |

### 4.2 Effect of Social Distancing

In Fig. 2, the trajectories of the exposed and infected subpopulations are shown as a function of time. By decreasing *σ* from 1 to 0.5, we can see the effect of flattening the curve as social distancing is increased. This is a natural consequence of the model because the effective contact rate is reduced (i.e. *σβ* is reduced). The base-case, in which the effective contact rate is unchanged (i.e. *σ* = 1), results in a peak population infection rate of 9% after just 44 days since the disease starts spreading. With *σ* = 0.75, the peak infection rate reduces to 6.3% after an additional 16 days, and *σ* = 0.5 results in an infection rate of only 2.6% nearly 46 days later. This shows that social distancing may play an important role, not only in reducing the prevalence of infection, but also in providing people more time to equip themselves with resources to combat the spread of the disease.

**Figure 2:**
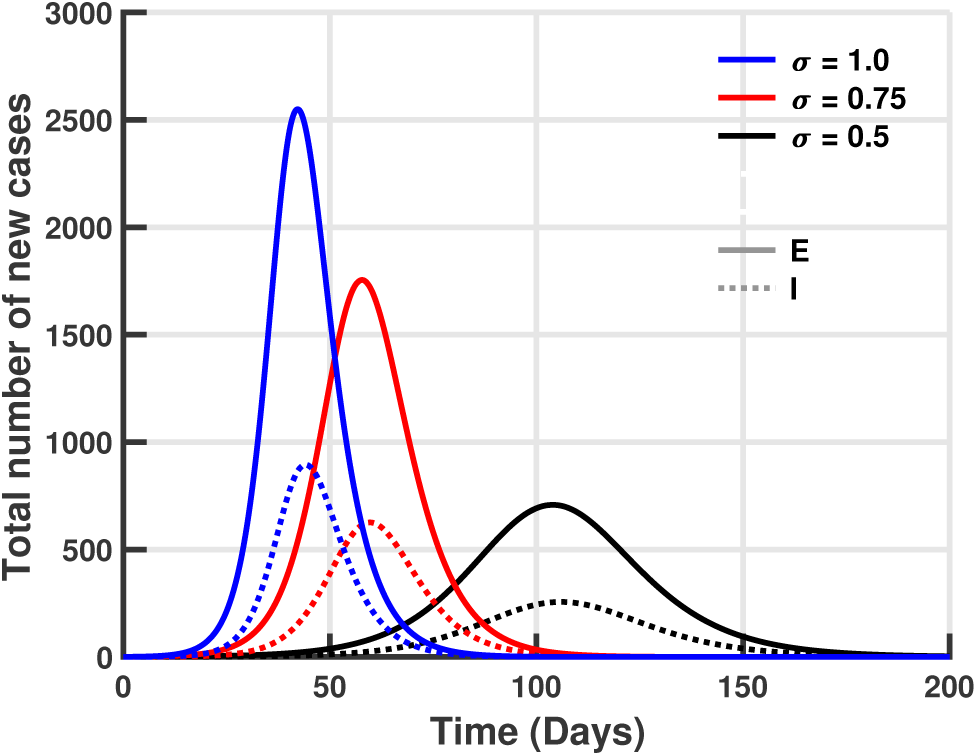
2019-nCoV SEIR Model with Social Distancing. The cases *σ* = 1.0, 0.75, 0.5 are shown respectively by the curves in blue, red, and black. Trajectories for the exposed population are solid curves and trajectories for the infected populations are dotted curves. The effect of social distancing on flattening the curve is seen clearly.

In Fig. 3, we see that for *σ ∈* [0, 0.3], there is a small number of new infections *T*_*N*_ in the population compared to the number of infections for *σ* > 0.3. In this region, *T*_*N*_ grows rapidly until the curve saturates (i.e. nearly the entire population is infected). Thus, the added value of enforcing social distancing for 100% of the population, corresponding to *σ* = 0, is insignificant when compared to the case *σ* = 0.3, where only 70% of the population obeys the protocol. Furthermore, there is only one new infection in the population when *σ* = 0.149, but nearly 90% of the population is infected when *σ* = 0.763. This behavior suggests that a recommendation for the value of *σ* would benefit from this analysis, by avoiding the extremes of the unit interval.

**Figure 3:**
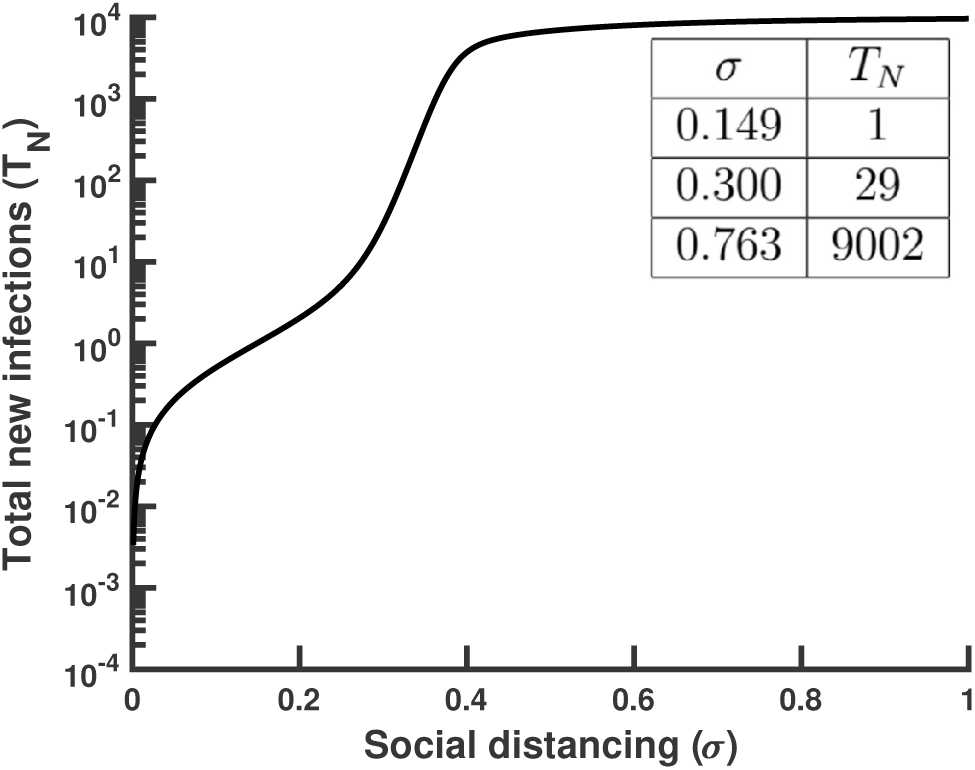
Total new infections as a function of social distancing. The curve exhibits interesting behavior for *σ ∈* [0.149, 0.763].

Thus, our numerical results suggest that there is a significant impact of social distancing on the outcome of a treatment program for this disease, thereby necessitating an optimal treatment protocol, which we discuss in the sections to follow.

## 5 Optimal Control

Our goal here is to find an optimal treatment strategy that will minimize the total disease burden, while minimizing the cost of implementing such a strategy.

### 5.1 Formulation

We introduce a time-dependent control *σ* = *u*(*t*), representing the effect of social distancing, in the model. This results in the following system:

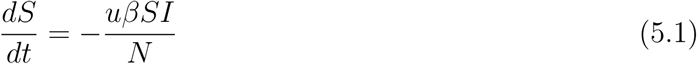

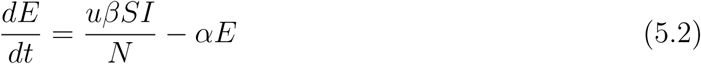

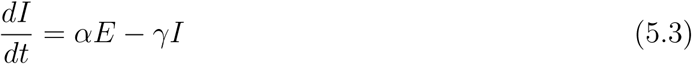

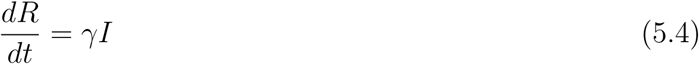

where *N*(*t*) =*S*(*t*)+*E*(*t*)+*I*(*t*)+*R*(*t*).

Note that for non-negative initial conditions and bounded Lebesgue-measurable controls, the state system admits non-negative bounded solutions [6]. The objective functional that we seek to minimize over a finite time horizon [0, *t*_*f*_] is:

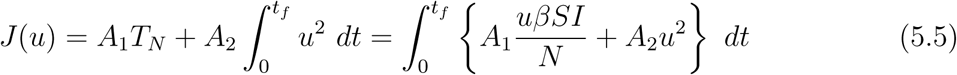

In (5.5), we assume the cost term is a quadratic function of the control [4, 6]. Here *A*_1_ represents the balancing factor associated with the total number of new infections *T*_*N*_. The balancing factor associated to the cost component *u*^2^ is denoted by *A*_2_.

We consider the set of admissible (bounded) control functions given by

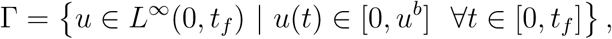

where *t*_*f*_ denotes the final time of the study period and *u* is Lebesgue-measurable with upper bound *u*^*b*^. Thus, we seek an optimal control *u*^*\**^ such that

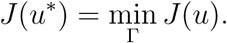

### 5.2 Optimality System

The existence of the optimal control follows from standard results due to the structure of the state system and the uniform *L*^*∞*^ bounds on the states and the controls [2]. To characterize the optimal control we use Pontryagin’s Maximum Principle [8], which allows us to utilize adjoint functions to transform the optimization problem into a problem of determining the pointwise minimum of the Hamiltonian, relative to *u*. The Hamiltonian is constructed from the functional (5.5), with the underlying state dynamics attached:

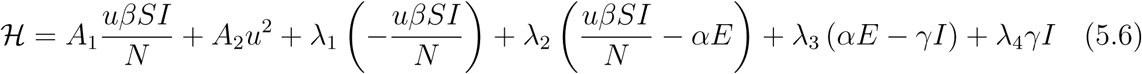

where *λ*_1_, *λ*_2_, *λ*_3_, *λ*_4_ are the adjoint variables associated to the states *S, E, I, R*.

For our system of ODEs, we are able to use Pontryagin’s Maximum Principle to obtain adjoint functions, which track the changes in our objective functional due to the state variables. The theorem below gives the ODE system and the boundary conditions for our adjoint functions and the corresponding characterization of an optimal control. The optimality system consists of our state and adjoint systems together with this control characterization, and we will solve this optimality system numerically to obtain the optimal control. The optimality system of equations results from taking the appropriate partial derivatives of (5.6) with respect to the associated state variable.

#### Theorem 5.2.1

*There exists an optimal control u*^*\**^ *and corresponding solution vector* (*S*^*\**^, *E*^*\**^, *I*^*\**^, *R*^*\**^) *that minimizes J* (*u*) *over* Γ. *Furthermore, there exist adjoint functions λ*_1_(*t*), *λ*_2_(*t*), *λ*_3_(*t*), *λ*_4_(*t*) *with transversality conditions*

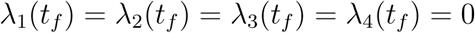

*Also, the following characterization holds:*

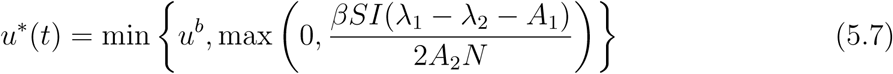

*Proof:* Corollary 4.1 of [2] gives the existence of an optimal control due to the convexity of the integrand of (5.5) with respect to the control *u, a priori* boundedness of the state solutions, and the *Lipschitz* property of the state system with respect to the state variables. Applying Pontryagin’s Maximum Principle, we obtain the adjoint system

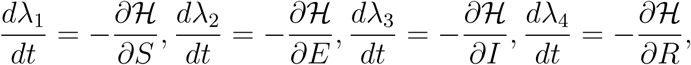

with zero final time conditions. To get the characterizations of the optimal control given by (5.7), we solve the equations on the interior of the control set 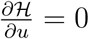. Using the bounds on the control, we obtain the desired characterization.

The optimality system consists of the state system with the initial time conditions, the costate (adjoint) system with the terminal time conditions, and the control characterization. Due to *a priori* boundedness of the state and adjoint systems, we obtain the uniqueness of the optimal control for small final time *t*_*f*_. This small time condition is due to opposite time orientations of the state system and the adjoint system [4].

For the sake of completeness, we provide the adjoint functions here:

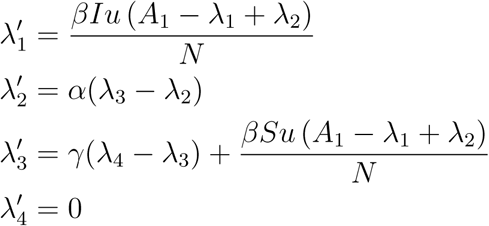

### 5.3 Method of Numerical Computations

We implement an iterative procedure to numerically solve the boundary value problem of the optimality system, consisting of eight ordinary differential equations comprising the state and adjoint equations, coupled with the control characterization. Numerical computation begins with an initial guess for the control and then uses a forward fourth-order Runge-Kutta scheme to solve the state equations over the time interval [0, *t*_*f*_] partitioned into *n* subintervals. Using the resulting state values and the given final time values, the adjoint system is then solved backward in time (due to the transversality conditions), again using a fourth-order Runge-Kutta method. Then, the control is updated by using a convex combination of the previous control and the value from the characterization. This iterative process continues until convergence, which is set to occur when the relative error between all state variables, the adjoint functions, and the control function is less than a specified value *δ*, i.e., when 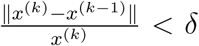, where ∥ *·*∥ is the *L*^*∞*^-norm. The method of numerical computation discussed above can be summarized as the algorithm below [4, 6]:

Here, 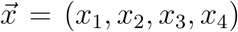 and 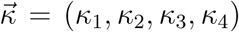 denote the vector approximations for the states and adjoints.

**Step 1:** Make an initial guess for *u* over the interval [0, *t*_*f*_].

**Step 2:** While (*δ*∥ *x*^(*k*)^∥ *−* ∥ *x*^(*k*)^ *− x*^(*k−*1)^∥ < 0), perform Steps 3 to 5.

**Step 3:** Solve 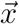 forward in time (using a fourth-order Runge-Kutta scheme) according to its system of differential equations in the optimality system.

**Step 4:** Using the transversality condition *κ*_4_ = *κ*(*t*_*f*_) = 0 and the stored values for *u*, 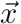, solve 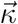 backward in time (using a fourth-order Runge-Kutta scheme) according to its system of differential equations in the optimality system.

**Step 5:** Update *u* by entering the new 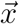 and 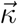 values into the characterization of the optimal control.

To obtain meaningful optimal control profiles, reasonable estimation of the balancing factors *A*_1_ and *A*_2_ is important. To estimate these balancing factors, we first compute the median value of *T*_*N*_ (*≈* 6, 816) from the region *σ∈* [0, 1]. We discretized *σ* into 1, 001 values, with median value 0.5. In the balanced situation, we have

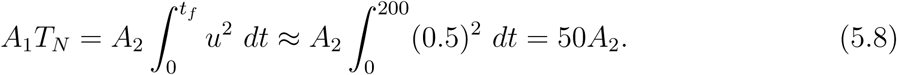

This implies *A*_2_*/A*_1_ = 136.32. Hence we take *A*_1_ = 1 and *A*_2_ = 136.32.

### 5.4 Numerical Results

We performed all numerical computations in *MATLAB R2019b* [10]. The value of *δ* was set at 10^*−*3^ for all computations. For the base-case (*A*_1_, *A*_2_) = (1, 136.32), we obtain an optimal control *u*^*\**^ that is identically zero for *t ∈* [0, *t*_*f*_]. (Fig. 4). From a mathematical point of view, this is perhaps not surprising since *T*_*N*_ is linear in the control and hence the objective functional *J* (*u*) is a multiple of *u*. From a control standpoint, it is apparent that *u*^*\**^*≡* 0 corresponds to the enforcement of social distancing in the entire population for the duration of the study. Clearly, this measure may be considered extreme, and in some cases even logistically infeasible. However, it is a necessary step for the purpose of minimizing infection risk.

**Figure 4:**
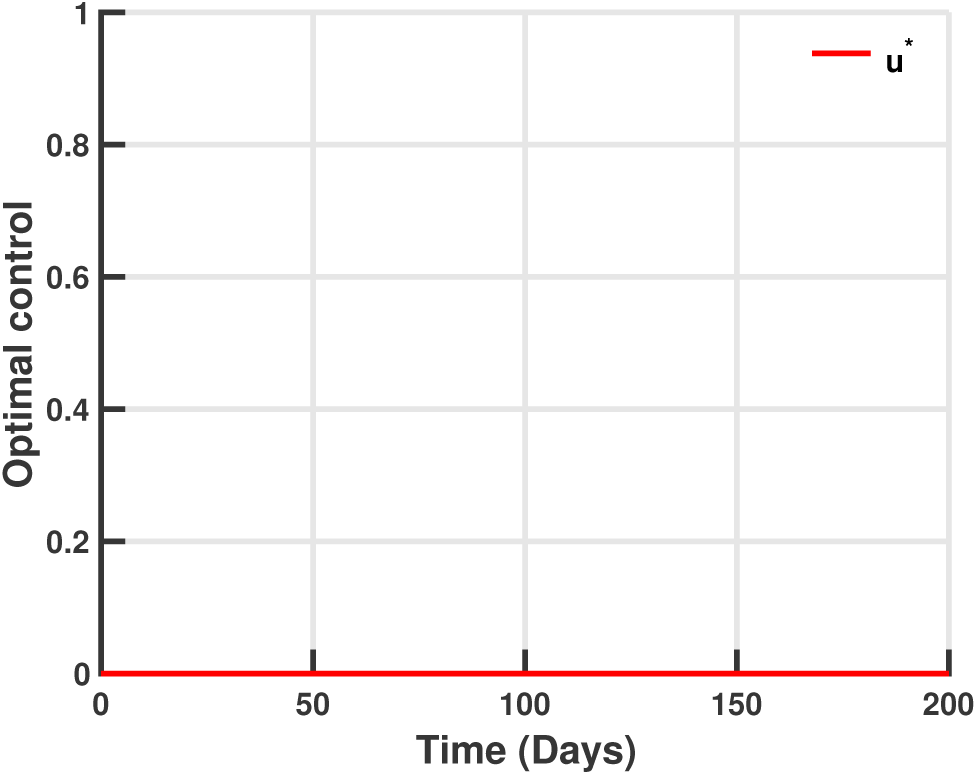
Effect of including social distancing in the optimal control. Optimal control solution profile *u*^*\**^ for *A*_1_ = *A*_2_ = 1.

## 6 Conclusion

Recent months have shown that 2019-nCoV poses a dangerous risk to humans. In this study, we developed a SEIR model that incorporates social distancing as a preventive measure for disease spread. We conducted analyses of the model, including formulation of the basic reproduction number and stability of the disease-free equilibrium (defined as a state where the disease is eradicated). Our analyses show that social distancing play directly affects the basic reproduction number, and hence the stability of the disease-free equilibrium. Our model simulations demonstrate that the total disease burden from 2019-nCoV can be mitigated by flattening the curve with a carefully chosen value for the coefficient of social distancing. Hence, we also formulated an optimal control problem and identified an optimal preventive measure for the population.

Our model has some limitations. In order to simplify the model, we have ignored the dynamics of age structure, disease mortality, and the presence of quarantined individuals. We assumed that all individuals in a given compartment are identically infectious, which might ignore potential effects caused due to population heterogeneity. Finally, further refinement of the objective functional is necessary in order to ensure a successful control protocol of 2019-nCoV.

## Data Availability

All relevant data are within the manuscript.

## Acknowledgement

The author would like to thank his advisor, Alan Hastings, for constructive suggestions that helped improve the paper.

